# Clinical and Immunohistochemical Determinants of Hepatocellular Carcinoma in Archival Liver Biopsies in Meru, Kenya

**DOI:** 10.64898/2026.02.21.26346789

**Authors:** Joshua Kibera, Jeff B. Bender, Francis M. Kobia, Rukia Kibaya, Milcom Gitonga, Frankline Gitonga, Francis Ondieki, Bactrin Kilingo, Stella Kepha, Mary Achakalong, Benti Gelalcha, Michael Mahero

## Abstract

**Background:** Hepatocellular carcinoma (HCC) is a leading cause of cancer-related death in sub-Saharan Africa (SSA). Differentiating primary HCC from metastatic liver tumors remains a significant diagnostic challenge. Understanding the prevalence and clinical predictors of HCC is crucial for improving diagnosis and patient care. This study examined the prevalence of hepatitis B virus (HBV), hepatitis C virus (HCV), and HCC, and clinical predictors of HCC.

**Methods:** We used immunohistochemical markers on archived liver tumor biopsies and analyzed the data using descriptive and logistic regression analysis.

**Results:** Among 58 liver carcinoma cases, 37.9% had HCC, and 62% had metastatic liver carcinoma (MLC). HCC was most common (61.5%) among middle-aged adults (50–59 years). HCC was more frequent in males (47.2%) than in females (22.7%). Over half of the patients (51.7%) tested positive for HBV. HCC was more prevalent in HBV-positive patients than HBV-negative ones (43.3% vs 32.1%). Hepatic fibrosis was identified in 27.6% of cases. HCC was more common in patients with fibrosis (56.2%) than in those without (31%). HCV infection was rare (6.9%) in this study. In multivariable logistic regression analysis, none of the examined predictors reached statistical significance (P>0.05). Patients aged 50–59 years, males, those with HBV infection, and hepatic fibrosis showed higher odds of HCC. Hepatocyte Paraffin-1 (Hep Par-1) demonstrated 97% specificity and a 95% positive predictive value (PPV) for differentiating HCC from MLC. The combined marker pattern of Hep Par-1 positive and AE1/AE3 negative was highly predictive of HCC (100% specificity, 100% PPV, and 93.2% diagnostic accuracy).

**Conclusions:** Our findings indicate that while the assessed risk factors tend to show directional association with HCC, as expected, larger studies are needed to determine their independent effects. The combined Hep Par-1 AE1/AE3 immunophenotype is more accurate than either marker alone. Therefore, this combined test is a valuable diagnostic tool for confirming HCC in resource-limited settings.

## 1. Introduction

Cancer is the third leading cause of death in Kenya, with an estimated 40,000 new cancer cases and 28,000 cancer deaths each year (although this is grossly underestimated) [1, 2].The Kenya Medical Research Institute documents that 80% of reported cases in the country are diagnosed at an advanced stage, leaving few options for survival [3, 4]. Hepatocellular carcinoma (HCC) is among the most prevalent types of cancer in Kenya and is ranked sixth in occurrence [1]. A striking difference of HCC from other cancers is that an underlying pathology is always present in the livers that develop primary cancer. In contrast to the Western countries, where lifestyle-related conditions like alcohol-linked liver cirrhosis, fatty liver, and hepatitis C have been identified as causal factors of HCC, while hepatitis B virus (HBV) and aflatoxin exposure have been identified as the major aetiological factors associated with HCC in sub-Saharan Africa (SSA) [5, 6]. Aflatoxins are fungal carcinogens associated with the *Aspergillus* fungi that are present in maize and are endemic in the East Africa region, as it presents the ideal environmental conditions for *Aspergillus*, particularly in arid and semi-arid areas [7–9]. Within Kenya, the eastern region is a drought-prone area that is globally recognized as an aflatoxin hotspot [10]. Studies in other regions of Africa have shown that aflatoxin consumption in individuals infected with HBV can facilitate mutations in the TP53 gene, dramatically increasing the risk of early HCC, even in the absence of cirrhosis [12].

The Meru region has been implicated as having a high cancer prevalence within Kenya [11]. HBV and Hepatitis C virus (HCV) are well-established risk factors for the development of HCC and are thought to be the main contributors to HCC in Africa [12, 13]. This study sought to determine the prevalence of HBV and Hepatitis C virus (HCV) in archival formalin-fixed paraffin-wax embedded (FFPE) liver biopsies diagnosed with HCC at a pathology laboratory in Meru. Investigating the prevalence of these viruses would provide much-needed context in the pathogenetic study of HCC and its association with other patient-level predictors.

Although PCR is highly sensitive for detecting viral DNA and RNA, including occult or low-level HBV and HCV infection [21], its availability in many developing countries remains limited. While PCR-based detection methods for viral DNA in FFPE have traditionally been preferred over other methods, they are expensive and require advanced molecular laboratory facilities and handling by skilled personnel familiar with DNA extraction from FFPE tissues [14, 15]. Although immunohistochemistry (IHC) is more readily available in many pathology laboratories and offers an alternative method for viral detection in low-resource settings, it is limited by its inability to detect quiescent HBV or HCV infection, which is characterised by the incorporation of viral DNA within hepatocytes without the expression of viral proteins [16]. It is worth noting that the expression of viral proteins, hepatitis B virus X protein (HBx), and middle surface proteins (LHBs/MHBs in HBV infection [17], and the core protein and non-structural proteins NS3, NS5A, and NS5B in HCV infection [18], are largely responsible for the oncogenic dysregulation of the hepatocyte cell cycle [19, 20]. Thus, these viral oncoproteins collectively contribute to the loss of normal cell-cycle control and hepatocarcinogenesis [18].

Unlike PCR, which detects viral nucleic acids regardless of activity, IHC highlights cells actively producing viral proteins [24]. IHC provides a practical complementary approach because it visualizes viral protein expression directly within hepatocytes, thereby identifying transcriptionally active infection with greater relevance to oncogenic pathways[22, 23]. In this study, we examined clinical predictors of HCC and also evaluated the diagnostic performance of immunohistochemical markers in distinguishing HCC from metastatic liver tumors.

## 2. Materials and methods

### 2.1. Immunohistochemical detection

In this study, all liver biopsy specimens received at Grace Park Pathology Services (now Aroha Cancer Center between 2017 and 2021 were retrieved and reviewed (n = 164). These stored liver tissues were obtained from cases with an abdominal mass based on ultrasound. Cases were excluded based on predefined criteria, including benign histological diagnoses, tissue blocks with limited tissue, necrotic or non-immunoreactive tissue, unavailable tissue blocks, or samples with missing key demographic data. After exclusions, we used 60 stored liver tissue samples from HCC cases (deemed to have appropriate quality tissue) to test for hepatitis-linked viral DNA and the presence of aflatoxin B1. Following this filtering process, 58 cases were deemed suitable for immunohistochemical analysis and included in the final study population. The variations in sample numbers reported at different stages of the study reflect this stepwise case selection process.

To protect the privacy of patients, new sections were cut from selected tissue blocks, and all slides were de-identified before analysis. De-identified slides were stored separately from original diagnostic materials to protect patient confidentiality and comply with ethical research standards. Ultrasound-guided biopsies from suspected cases were taken and further classified as HCC cases based on histopathological examination. A protocol previously used by Murugavel et al, 2007 [25] was employed to prepare archived tissues, HCV, and HBV markers identified using commercial assays (Dako Corp). Briefly, the paraffin sections were dewaxed in xylene and rehydrated through descending grades of alcohol. They were then washed in 0.5 M glycine in PBS (pH 7.2) to inhibit/remove fixing agents, such as aldehydes. Endogenous peroxidase activity was blocked with a fresh 0.3–0.5% solution of H_2_O_2_ in absolute methanol for 30 min. The slides were then washed in Tris buffer for 5 min and incubated in a few drops of 3% normal goat serum for 20–30 min. Slides with representative sections were subjected to immunohistochemistry with monoclonal antibody Hep Par 1, AE1/AE3, anti-HCV, anti-HBV, and GPC-3 (Dako Corp) using the avidin biotin technique with a primary antibody dilution of 1:40. Adjacent non-tumorous hepatocytes were taken as positive controls as previously described [16].

The immunohistochemical staining result was independently interpreted by two pathologists. Given that HBV and HCV IHC staining are non-standard, both reviewers first familiarized themselves with established staining patterns. HBV positivity was defined by the presence of combined nuclear and cytoplasmic staining. HCV positivity was identified by staining within hepatocytes and stromal cells. Independent scoring results were compiled, and cases with discrepant interpretations were reviewed jointly until consensus was reached, ensuring consistency and diagnostic reliability.

### 2.2. Data management

Cases with a histological diagnosis of either hepatocellular carcinoma (HCC) or metastatic carcinoma to the liver were included. The variables included in the analysis included patient age category, sex, fibrosis status (Yes/No), immunohistochemical marker results for Hepatocyte Paraffin 1 (Hep Par-1) and AE1/AE3 cytokeratin cocktail, HBV and HCV status (Positive/Negative/CBD (uncertain), and staining patterns.

We defined the primary dependent variable, Diagnosis_bin, as 1 for HCC and 0 for metastatic carcinoma. Age was categorized into <50, 50-59, 60-69, and ≥70 years. Sex was ≥ coded as male = 1 and female = 0. Fibrosis was coded as binary (Yes = 1, No = 0). HBV and HCV status were initially coded as Positive, Negative, CBD–uncertain). The “CBD” values were treated as missing after sensitivity analyses showed no effect on model estimates. Immunohistochemical variables HepPar1_bin and AE1AE3_bin were binary (Positive = 1, Negative = 0). A composite marker, Marker Pattern, was defined as 1 if HepPar-1 = 1 AND AE1/AE3 = 0, and 0 otherwise.

### 2.3. Statistical analyses

We used descriptive statistics to summarize categorical variables as frequencies (n) and percentages (%) for HCC versus metastatic carcinoma groups. Univariate comparisons used chi-square or Fisher’s exact tests as appropriate, with p-value <0.05 denoting statistical significance.

Logistic regression models were fitted to estimate adjusted odds ratios (ORs) and 95% confidence intervals (CIs) for predictors of HCC versus metastasis. The multivariable model included age category, sex, fibrosis status, HBV status, and HCV status. Sensitivity analyses addressed the “CBD” (uncertain) category by treating it first as positive, then as negative; identical results permitted treating CBD values as missing for the final model. Odds ratios were calculated by exponentiating regression coefficients. The model fit was assessed via pseudo-R² and the likelihood-ratio test.

For the histopathologic marker analysis, a logistic model was attempted, including HepPar1_bin, AE1AE3_bin, sex, and age category. However, quasi-separation (perfect prediction) was found, prompting us to use a diagnostic performance approach instead. For each marker and for the composite marker pattern, sensitivity, specificity, positive predictive value (PPV), negative predictive value (NPV), accuracy, and counts (TP, FP, FN, TN) were calculated by 2×2 cross-tabulation of marker result vs outcome (HCC vs metastasis). All statistical tests were two-sided, and all code and data manipulations were reviewed for reproducibility. All statistical analyses were conducted using Python 3.11 in a Jupyter Notebook environment. Data management and descriptive statistics were performed using the pandas library, and inferential analyses (chi-square tests, logistic regression models) were conducted using the statsmodels package. A p-value of < 0.05 was considered statistically significant, and 95% CIs were reported where appropriate.

## 3. Results

### 3.1. Clinical predictors and their association with HCC

Of the 58 cases examined, 37.9% (n=22) were histologically confirmed as HCC, and 62% (n=36) were metastatic liver carcinoma (MLC). A higher proportion (32.8%) of patients (both HCC and MLC) were younger than 50 years, the least proportion (17.2%) was 70 years and above. Males constituted 62.1% of the study population. The proportion of HCC cases was highest (61.5%) in the 50–59 years age group, and least (20%) among those aged 70 years. ≥ HCC cases were more frequent in males (47.2%) than in females (22.7%), while metastatic cases were more frequent in females (77.3%). Metastatic cases were most frequent (80%) in older patients (≥70 years). Hepatitis B surface antigen was positive in 51.7% of cases, with HCC ≥ detected in 43.3% of HBV-positive patients. Hepatitis C infection was less common (6.9%) in this study. Fibrosis was detected in 27.6% of cases. HCC was detected more frequently among individuals with fibrosis (56.2%) compared to those without (31.0%) (Table 1).

**Table 1.** Logistic regression analysis of factors associated with hepatocellular carcinoma (HCC) vs. metastatic liver carcinoma (MLC) (N = 58)

| Variable | Category | HCC<br>n (%) | MLC<br>n (%) | Total<br>n (%) | p-value | Adjusted OR (95%<br>CI) |
| --- | --- | --- | --- | --- | --- | --- |
| **Age group** | **<50 Y** | 8 (42.1) | 11 (57.9) | 19 (32.8) | — | **Ref** |
|  | 50–59 Y | 8 (61.5) | 5 (38.5) | 13 (22.4) | 0.247 | 2.58 (0.52–12.83) |
|  | 60–69 Y | 4 (25.0) | 12 (75.0) | 16 (27.6) | 0.329 | 0.46 (0.09–2.21) |
| | $\geq 70$ Y | 2 (20.0) | 8 (80.0) | 10 (17.2) | 0.24 | 0.32 (0.05–2.13) |
| **Sex** | Female | 5 (22.7) | 17 (77.3) | 22 (37.9) | — | **Ref** |
|  | Male | 17 (47.2) | 19 (52.8) | 36 (62.1) | 0.127 | 2.75 (0.75–10.10) |
| **Hepatitis B** | Negative | 9 (32.1) | 19 (67.9) | 28 (48.3) | — | **Ref** |
|  | Positive | 13 (43.3) | 17 (56.7) | 30 (51.7) | 0.163 | 2.54 (0.69–9.38) |
| **Hepatitis C** | Negative | 21 (38.9) | 33 (61.1) | 54 (93.1) | — | **Ref** |
|  | Positive | 1 (25.0) | 3 (75.0) | 4 (6.9) | 0.438 | 0.35 (0.02–5.03) |
| **Fibrosis** | No | 13 (31.0) | 29 (69.0) | 42 (72.4) | — | **Ref** |
|  | Yes | 9 (56.2) | 7 (43.8) | 16 (27.6) | 0.274 | 2.17 (0.54–8.63) |

Multivariable logistic regression analysis was done to identify factors independently associated with HCC, using metastatic carcinoma as the reference category (Table 1). Male sex, fibrosis, and hepatitis B positivity were positively associated with HCC. But hepatitis C infection and older age groups showed an inverse relationship with HCC. However, the regression model documented that none of the predictors mentioned above reached statistical significance. Specifically, patients aged 50–59 years had higher odds of HCC compared to the rest of the age category, but the difference was not statistically significant (p=0.247; OR = 2.5, 95% CI: 0.52–12.83). Interestingly, individuals aged ≥70 years had lower odds of HCC compared to those aged <50 years (p = 0.24; OR = 0.32, 95% CI: 0.05–2.13). Males had higher odds of HCC than females (OR = 2.75, 95% CI: 0.75–10.10). Similarly, patients with HBV infection also showed increased odds of HCC (p=0.163; OR = 2.54, 95% CI: 0.69–9.38). Patients with fibrosis had higher odds of HCC, and again the difference was not statistically significant (p = 0.274; OR = 2.17, 95% CI: 0.54–8.63).

We also carried out an analysis of whether HBV infection increases the odds of fibrosis. Fibrosis was observed more frequently in HBV-negative (32.1%) patients than in HBV-positive (23.3%) ones. HBV infection was associated with decreased odds of fibrosis, although the result is not statistically significant (OR = 0.64; p = 0.56) (Table 2).

**Table 2.** Association Between Hepatitis B Status and Fibrosis in the Study Population.

| Hepatitis B Status | No Fibrosis | Fibrosis Present | Total |
| --- | --- | --- | --- |
| HBV Positive | 23 (76.7%) | 7 (23.3%) | 30 |
| HBV Negative | 19 (67.9%) | 9 (32.1%) | 28 |
| Total | 42 | 16 | 58 |

OR = 0.64, 95% CI: 0.20–2.05, Fisher’s Exact $p$ -value = 0.56)

We also tested whether fibrosis is a predictor of HCC (independent of HBV status) and found that most patients with fibrosis (56.2%) had HCC. Patients with fibrosis had nearly 3-fold (OR=2.9) higher odds of HCC than those without, but this association was not statistically significant in our dataset (p-value = 0.13) (Table 3).

**Table 3.** Association Between Fibrosis Status and HCC in the Study Population.

| Fibrosis | MLC n (%) | HCC n(%) | Total |
| --- | --- | --- | --- |
| Present | 7 (43.8%) | 9 (56.2%) | 16 |
| Absent | 29 (69.0%) | 13 (31.0%) | 42 |
| Total | 36 | 22 | 58 |

OR = 2.87; 95% CI = 0.88 – 9.38; Fisher’s Exact $p$ -value = 0.13

### 3.2. Distribution patterns of immunohistochemical markers in HCC and MLC

Distribution of composite staining patterns of Hep Par-1 and AE1/AE3 among liver carcinoma shows Hep Par-1+/AE1-AE3 phenotype is restricted to HCC (42.9%), indicating its diagnostic specificity (no metastatic cases detected in this pattern). However, the inverse phenotype was dominant (86.5%) among MLC (Table 4).

**Table 4.**
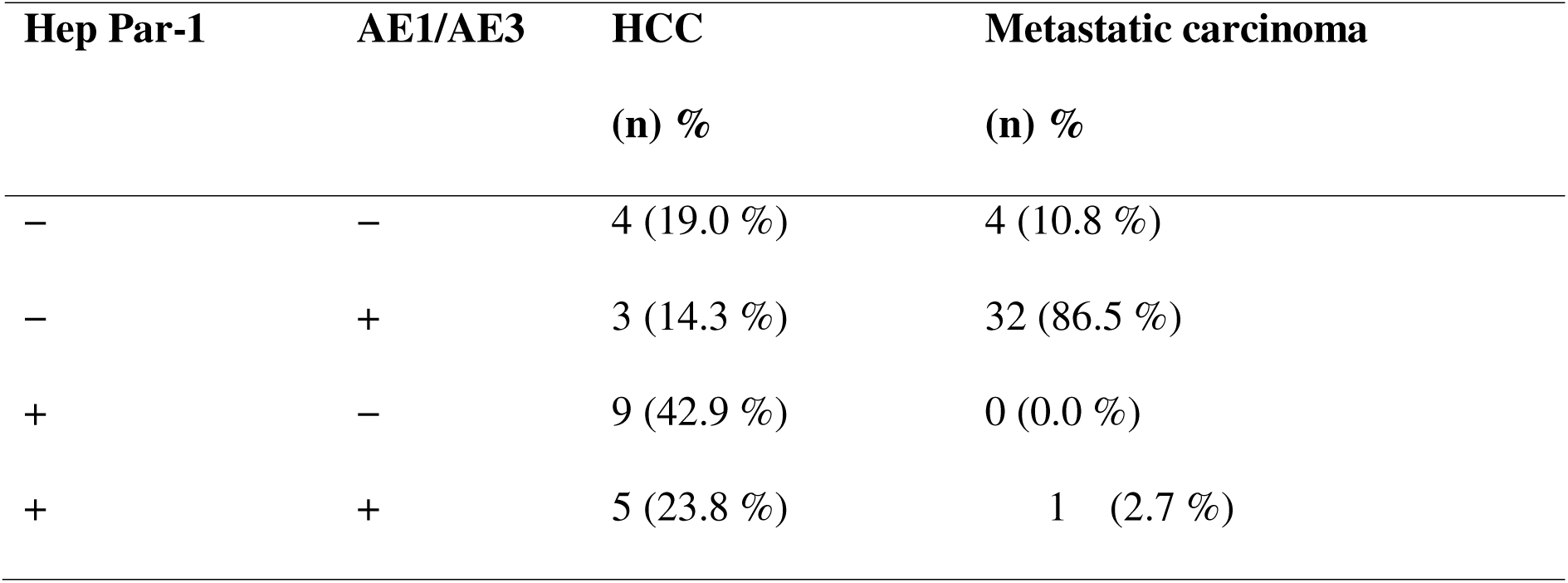
Distribution of Hep Par-1 and AE1/AE3 co-expression patterns in hepatocellular carcinoma (HCC) and metastatic liver carcinoma (MLC)

### 3.3. Diagnostic performance of individual immunohistochemical markers

When assessing individual immunohistochemical markers, Hep Par-1 demonstrated good diagnostic performance for HCC, with a sensitivity of 72% and a specificity of 97%. The PPV was high (95%), indicating that a positive Hep Par-1 stain serves as a strong rule-in test for HCC. In contrast, AE1/AE3 showed poor discriminatory power in identifying HCC from metastatic carcinoma in this dataset (Table 5).

**Table 5.**
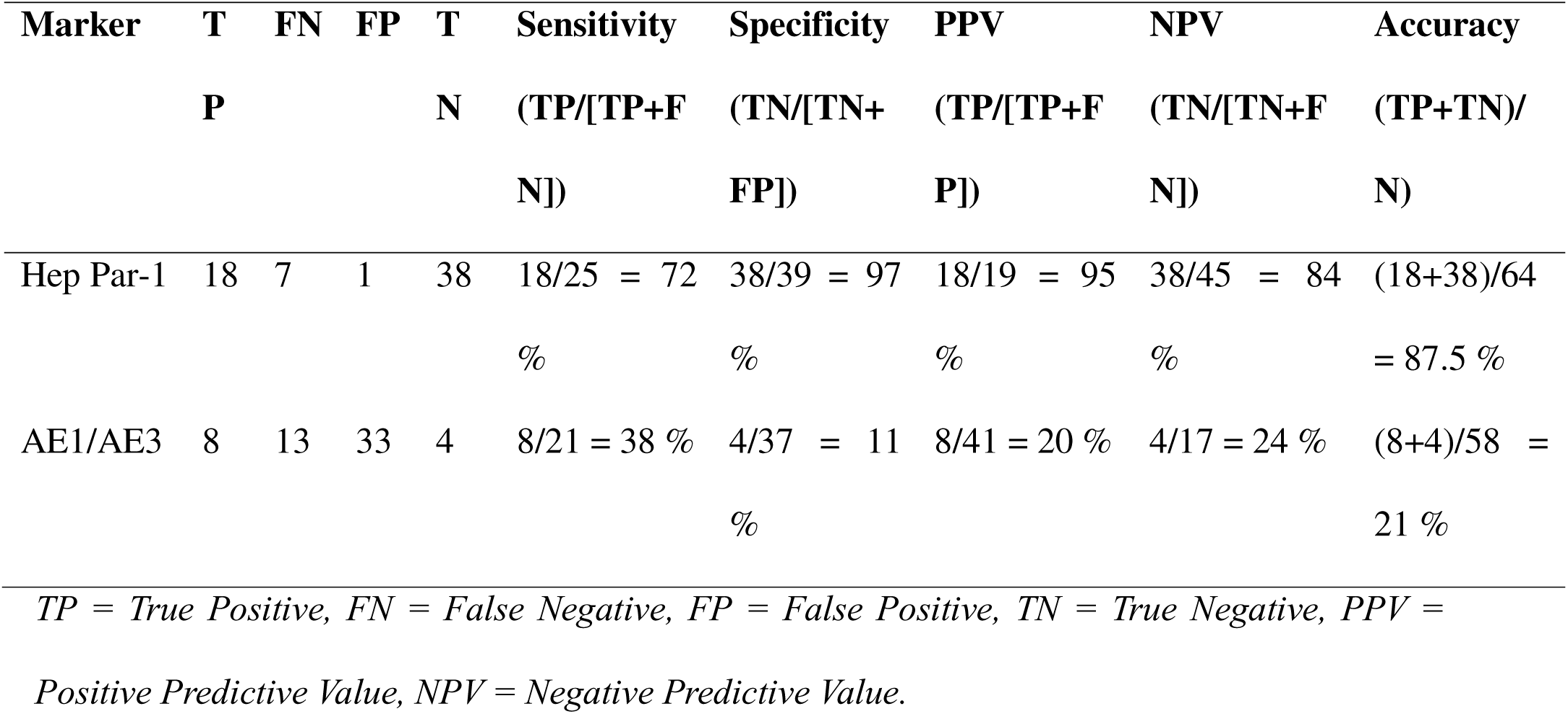
Diagnostic performance of histopathologic markers in distinguishing hepatocellular carcinoma (HCC) from metastatic liver carcinoma (MLC)

| Marker | T | FN | FP | T | Sensitivity | Specificity | PPV | NPV | Accuracy |
| --- | --- | --- | --- | --- | --- | --- | --- | --- | --- |
|  | P |  |  | N | (TP/[TP+F<br>N]) | (TN/[TN+<br>FP]) | (TP/[TP+F<br>P]) | (TN/[TN+F<br>N]) | (TP+TN)/<br>N) |
| Hep Par-1 | 18 | 7 | 1 | 38 | 18/25 = 72<br>% | 38/39 = 97<br>% | 18/19 = 95<br>% | 38/45 = 84<br>% | (18+38)/64<br>= 87.5 % |
| AE1/AE3 | 8 | 13 | 33 | 4 | 8/21 = 38 % | 4/37 = 11<br>% | 8/41 = 20 % | 4/17 = 24 % | (8+4)/58 =<br>21 % |

*TP = True Positive, FN = False Negative, FP = False Positive, TN = True Negative, PPV = Positive Predictive Value, NPV = Negative Predictive Value.*

### 3.4. Diagnostic performance of the combined immunophenotypic signature

The combined immunophenotypic signature (Hep Par-1 and AE1/AE3) demonstrated excellent diagnostic ability in differentiating HCC from metastatic carcinoma. The Hep Par-1 / AE1-AE3 phenotype correctly identified 9 of 12 HCC cases (true positives) and showed no false positives, with a sensitivity of 75.0% and a PPV of 100%. Although this composite pattern will miss a significant proportion of HCC cases (those without this classical signature), its high specificity makes this pattern a highly reliable rule-in indicator of HCC, particularly in challenging biopsy cases.

_Conversely, the Hep Par-1_ / AE1-AE3 metastatic phenotype correctly classified all 32 metastatic cases (true negatives), with no misclassification, resulting in a 100% specificity and an NPV of 91.4%. Overall diagnostic accuracy among cases with decisive immunophenotypes was 93.2% (41/44) (Table 6).

**Table 6.**
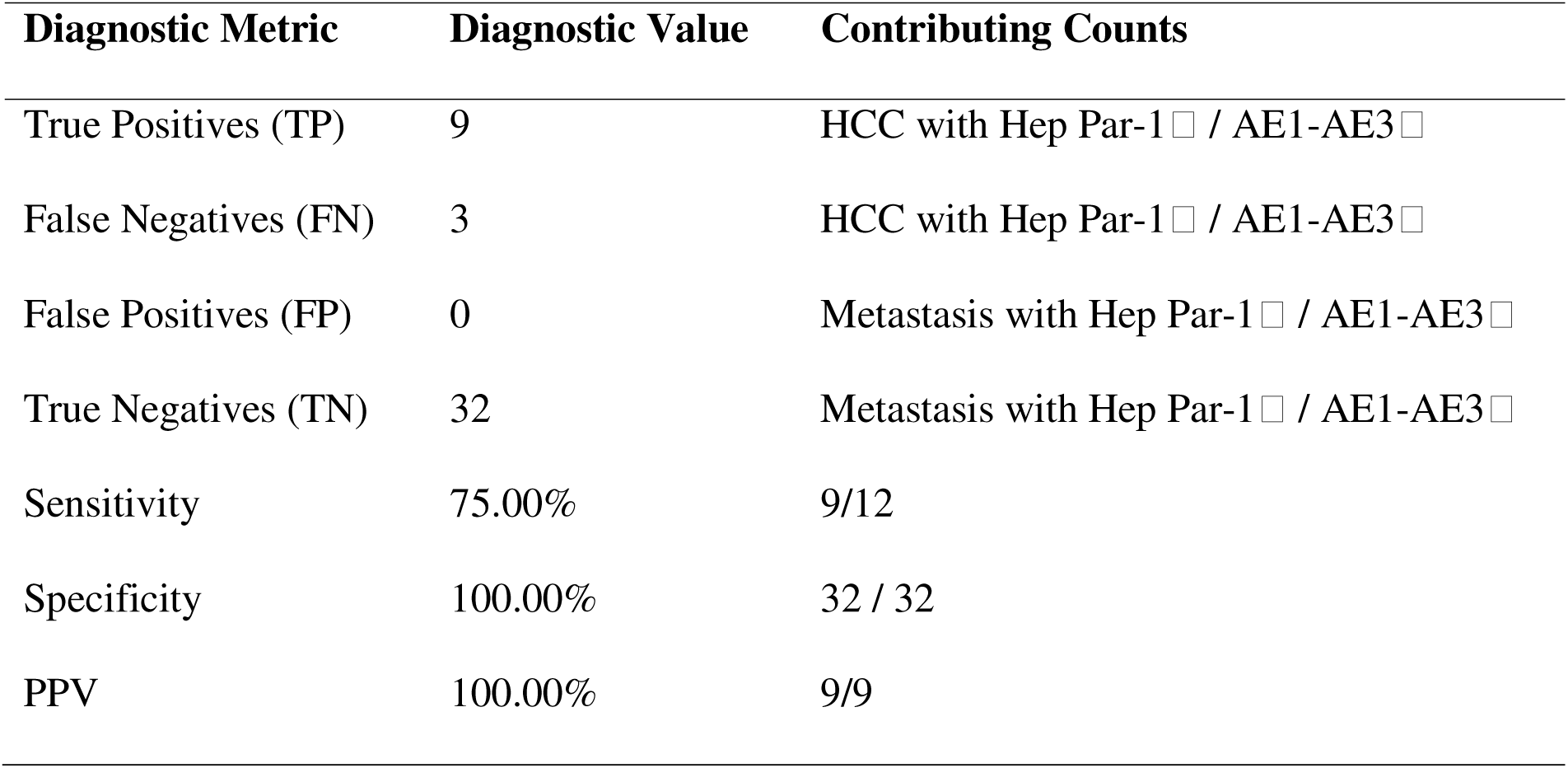
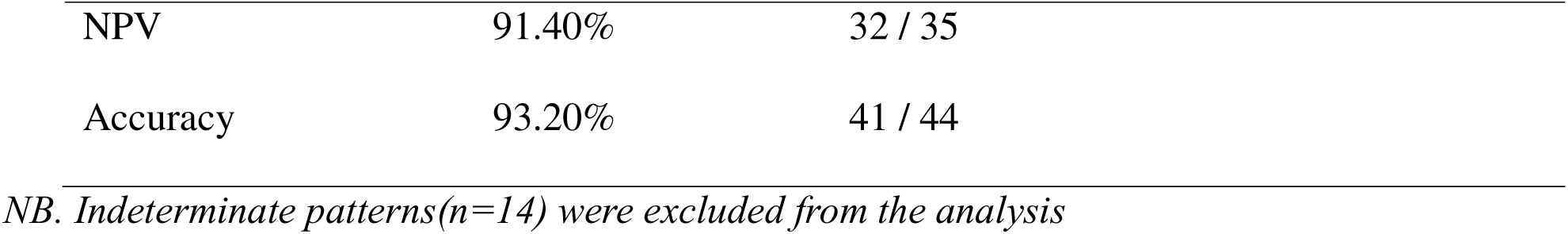
Diagnostic performance of the combined Hep Par-1 / AE1-AE3 signature for distinguishing HCC from metastatic carcinoma.

## 4. Discussion

In this study, we analyzed clinical predictors and immunohistochemical markers to distinguish HCC from metastatic carcinomas in the liver. Our multivariable logistic regression showed that male sex, hepatic fibrosis, and HBV positivity were associated with increased odds of HCC compared to metastatic disease. Although none reached traditional statistical significance, these associations are biologically plausible and align well with established epidemiological evidence [26–31]. The lack of a significant association between HBV/HCV and HCC in our study also suggests that other factors, such as aflatoxin exposure, alcohol-related liver disease, chronic environmental toxin exposure, and cirrhosis from mixed etiologies, may be contributing to HCC occurrence [32, 33]. Thus, the results of our study open a reasonable avenue to investigate these other potential factors, especially aflatoxin exposure, given the regional context [34, 35].

HCC is well-known to occur more commonly in men and in the setting of chronic liver disease with fibrosis or viral hepatitis [27]. The frequent occurrence of HCC in males has been reported in a large prospective cohort study, with sex-adjusted estimates showing over a 4-fold increase in men [26]. Studies showed that this gender difference is not only due to behavioral factors but also due to a complex interplay between sex hormones and viral pathogenesis (androgens promoting HBV replication, whereas estrogens offer protective effects) [27, 36]. The wide confidence intervals and non-significant p-values in this study likely reflect our limited sample size (lack of power) rather than the absence of effect.

In this study, age displayed an interesting non linear pattern. The increased odds in the 50-59 years age group (OR =2.58) compared to <50 (reference group), followed by a decline in older age categories (OR =0.46 for 60-69, and 0.32 for ≥70). Previous works have consistently ≥ reported older age as a risk factor for HCC [28, 37]. In this study, though we believe the lack of statistical association between older age and HCC is likely due to sample size, the established relationship might have been modified by selection bias (differences in referral or biopsy sampling). In addition, survivor bias in older patients may also contribute to this variation, as older patients may have higher competing risks from non-HCC mortality, limiting HCC detection [28].

The positive associations observed between HCC and HBV positivity and hepatic fibrosis are consistent with established molecular mechanisms. HBV can increase the risk of HCC in several ways, including direct insertional mutagenesis into host DNA [38], HBx protein-mediated transactivation of oncogenic pathways and genomic instability induced by chronic inflammation [17, 29, 30]. Previous studies showed a 5-to 50-fold increase in the risk of HCC in HBV seropositivity [28, 31, 39–41].

Hepatic fibrosis, particularly the advanced type, and cirrhosis, represent one of the most important risk factors for HCC. The direction of the effect in our study is in line with previous studies showing cirrhosis increases HCC incidence about 42-fold compared to non-cirrhotic subjects [1], as chronic hepatocellular injury leads to cycles of necrosis and regeneration, which promotes mutation accumulation and chromosomal instability, and then HCC [26, 42, 43].

In this study, patients with HBV infection did not show increased odds of fibrosis, and the result showed fibrosis was slightly less frequent in HBV-positive cases, though the difference was not significant. However, the observed direction of effect in this study is not consistent with previous findings that demonstrated a very strong association between HBV infection and hepatic fibrosis [41, 44–46]. This absence of association in our study could be due to fibrosis often depends on the duration and severity of disease, which our dataset does not capture [45, 47]. In addition, fibrosis in our dataset may be due to chronic liver damage that may come from multiple other causes, including chronic viral infection, steatohepatitis (alcoholic and non-alcoholic), or other liver injury processes, and not merely HBV infection [48, 49].

In contrast, fibrosis demonstrated a notable increase with HCC, though the association was not statistically significant (p=0.13). The observed trend (effect direction) toward a higher frequency of HCC among patients with fibrosis (OR = 2.87) is consistent with the established biological pathway in which chronic liver injury leads to fibrotic remodeling and subsequent hepatocarcinogenesis [40, 42, 43]. Absence of statistical significance is likely due to the few documented cases of fibrosis (n=16), rather than the absence of a true association. These findings reinforce the clinical importance of fibrosis surveillance and its role as an intermediate state in the liver cancer developmental continuum.

Hepatocyte paraffin 1 (Hep Par-1) immunohistochemistry showed a high degree of specificity (97%) for hepatocellular differentiation of HCC. Several studies have demonstrated that Hep Par 1 is one of the most reliable markers for confirming and differentiating HCC from other types of adenocarcinomas metastatic to the liver [50–56]. For instance, Ramadhani *et al* [52] reported Hep Par 1 has high diagnostic accuracy, with almost 100% sensitivity and specificity to diagnose HCC; Lugli *et al*. [50] reported that increased Hep Par 1 expression was found most frequently in HCC than in other carcinomas metastatic to the liver; Lau *et al*.[53], reported Hep Par 1 was sensitive (90%) and had good specificity for HCC.

As discussed above, most of the studies showed HepPar-1 as a sensitive and specific immunohistochemical marker for primary HCC. However, a few studies reported a very low sensitivity and specificity, particularly in cancer patients with morphologically poorly differentiated HCCs (difficult to identify from metastatic carcinoma) [56–59]. These same studies recommend the use of HepPar-1 in a panel with other markers.

In our study, AE1/AE3 positivity favors metastatic carcinoma and poorly identifies HCC, which is in line with previous studies [60]. However, some studies cautioned that AE1/AE3 can be seen in poorly differentiated HCC, suggesting the need for combination tests or a panel-based approach for accurate diagnosis of HCC and metastatic carcinoma, as each marker alone (HepPar-1 and AE1/AE3) has limitations [57, 61].

The combined use of Hep Par-1 and AE1/AE3 in this study provides a clinically reliable diagnostic tool for distinguishing HCC from metastatic carcinoma in liver biopsies. In this study, when combined, the Hep Par-1 / AE1-AE3 pattern becomes highly specific for HCC. It correctly identified all metastatic cases and all HCC cases, providing 100% specificity and 100% PPV. Previous studies also confirm that HCC is negative for cytokeratin AE1/AE3 due to its unique hepatocellular origin and cytokeratin profile [57, 61–63].

On the other hand, the Hep Par-1 / AE1-AE3 pattern strongly supported a diagnosis of metastatic carcinoma, accurately classifying all such cases (n=32) in the cohort. This is in line with a previous study showing that most metastatic carcinomas to the liver are AE1/AE3 positive and negative for the hepatocyte-specific marker, HepPar-1, [50, 57, 64]. This bidirectional reliability improves diagnostic confidence in routine practice, especially in small biopsies or cases where morphology alone is inadequate for definitive diagnosis.

Importantly, 24% of cases in this study showed indeterminate patterns (both markers positive or both negative). This indicates the need for careful integration of morphology, clinical history, and additional immunohistochemical markers when needed [65]. However, for cases with distinct patterns, the combined signature had an overall accuracy of 93.2%, outperforming the individual markers. Generally, these findings highlight Hep Par-1 as a key confirmatory marker for HCC. The combined Hep Par-1 / AE1/AE3 signature provides more reliable diagnostic confidence in differentiating HCC from metastatic carcinoma than using single immunophenotypic markers in isolation.

Misdiagnosis contributes to poor cancer outcomes in sub-Saharan Africa, where access to molecular diagnostics is limited, and cancer registries remain incomplete [66, 67]. Inaccurate distinction between HCC and metastatic liver tumors may lead to incorrect treatment decisions, missed opportunities for surveillance, or failure to identify underlying risk factors [67, 68]. Strengthening diagnostic capacity through validated, relatively low-cost tools such as immunohistochemistry is an important step toward improving cancer care and also enables correct epidemiologic mapping of HCC burden in the region [68].

Although the study demonstrates the feasibility of immunohistochemical detection of HBV and HCV in HCC tissue, the relatively small sample size and limited availability of clinical data constrain broader etiological interpretation. Correlation with serological markers for HBV and HCV, as well as exposure data for other known risk factors such as aflatoxin, was not consistently available. These limitations highlight the likelihood that additional etiological factors contribute to HCC in this population beyond viral hepatitis alone.

## 5. Conclusion

In this study, we assessed clinical predictors and immunohistochemical markers to distinguish HCC from MLC within a diagnostic setting. Although none of the assessed clinical factors (age, sex, hepatitis B or C infection, or fibrosis) reached statistical significance in the models, most of these predictors showed biologically consistent trends. Male sex, hepatitis B infection, and the presence of fibrosis were all associated with higher odds of HCC, and patients with fibrosis were nearly three times more likely to have HCC compared to those without. These patterns, although not statistically conclusive, reflect well-established hepatocarcinogenic pathways and likely indicate limited study power rather than the absence of true associations.

The immunohistochemical analysis yielded clearer diagnostic value. Hep Par-1 demonstrated excellent specificity and a high PPV for HCC, further strengthening its role as a confirmatory marker. AE1/AE3 alone provided limited discriminatory utility. However, when interpreted together, the composite signature of Hep Par-1 positivity with AE1/AE3 negativity was highly specific, correctly identifying all metastatic carcinomas and providing strong diagnostic confidence for primary hepatocellular origin. This combined pattern offers a simple, low-cost, and clinically meaningful approach that can help pathologists resolve challenging or morphologically vague liver lesions, especially in resource-limited settings. In addition, this study also highlights the need for integrated studies of HCC etiology in rural African settings that combine histopathology, clinical data, viral serology, and assessment of environmental carcinogens, particularly aflatoxin. The lack of validated tests for detecting aflatoxin-related DNA adducts shows the importance of developing alternative approaches to better understand HCC pathogenesis in Kenya and similar contexts where aflatoxin is prevalent.

## Data Availability

All data produced in the present study are available upon reasonable request to the authors.

## 6. Author contributions

Joshua Kibera, Jeff B. Bender, Francis M. Kobia, Stella Kepha and Michael Mahero conceived the study, wrote the original manuscript, and reviewed and edited the manuscript. Rukia Kibaya, Milcom Gitonga, Frankline Gitonga, and Mary Achakalonga conducted laboratory experiments (immunohistochemistry), and acquired and curated data. Francis Ondieki and Bactrin Killingo curated data, managed the study, and provided resources. Benti D Gelalcha analyzed the data, interpreted the results, and wrote original manuscript. All authors reviewed the final manuscript.

## 7. Ethics approval and consent to participate

Ethical approval for this study was granted by the ethics review committee of Mount Kenya University (approval number: 1264). Since this study used archival de-identified samples, consenting was not required.

## 8. Data statement

The data underlying this study are not openly accessible because of patient confidentiality but are available from the corresponding author upon request with reasonable notice.

## 9. Acknowledgments

The authors gratefully acknowledge the CGHSR UMN for supporting this study and the BDS UTK for covering the article processing fees.

